# Assessment of Accessibility, Availability, and Need of Cardiac Care at Primary Health Care Centres in Vadodara District of India: The CardioGram Pilot Project

**DOI:** 10.1101/2024.05.07.24307008

**Authors:** Shirish Rao, Ujjaini Rudra, Anoushka Arora, Sumanta Majumdar, Murtaza Gandhi, Naitica Darooka, Zahra Motiwalla, Sucharu Asri, Urmil Shah, Devarsh Shah, Dhrumil Patil, Eesha Shah, Katya Saksena, Priyansh Shah, Ami Bhatt, Siddhesh Zadey

**Author notes:** **<u>Correspondence:</u>** Shirish Rao, Seth G.S. Medical College and K.E.M Hospital, Mumbai, Maharashtra, India - 400012.

## Abstract

**Background:** There has been an increase in the burden of Cardiovascular Diseases (CVD), especially in rural India. Integrating screening and treatment for CVDs at the primary healthcare level has now become a necessity. Hence, this study was conducted to assess the accessibility, availability, and need for cardiac care with a special focus on ECG at Primary Healthcare Centers (PHC) in the Vadodara district of Gujarat.

**Methods:** A cross-sectional pilot study was carried out in 34 PHCs of the Vadodara District of Gujarat, India between January to March 2022. Data regarding the accessibility of PHC, distance from the nearest Tertiary Health Centre (THC), availability of ECG, drugs, treatment protocols, competency of staff, and burden of CVD, hypertension, and diabetes was collected by interviewing the medical officer of the PHC. Distances were measured in kilometers (km) and Travel times were measured in minutes (min). Descriptive analysis was performed using MS Excel.

**Results:** The average distance to reach a PHC, a Tertiary Care Center, and a Cardiac Care Center (CCC) is 14.8km, 37.1 km, and 55.5km, respectively, which accounts for 22.59 minutes, 62.3 minutes, and 85.5 minutes. Moreover, only 58% of the surveyed PHCs have the availability of ECGs, with a lesser percentage of healthcare workers(HCWs) who knew how to operate and/or read an ECG. 44.11% of the surveyed PHCs had functional ECGs and employed them where indicated. Most of the CVD cases were referred to higher centers. Six PHCs had an urgent need for ECG deployment.

**Conclusion:** Accessibility and availability of cardiac care services, especially ECGs is poor in PHCs of Vadodara district. There is an urgent need not only for services but also for efficient training of medical officers for performing, interpreting as well as managing cases of acute myocardial infarction.

## Introduction

The cardiovascular disease (CVD) accounts for nearly a quarter (24.8%) of all deaths in India. The burden of CVD-causing risk factors such as the use of tobacco is rising and the prevalent cases of CVD are likely to increase substantially[1,2]. Incidence of CVD and CVD risk factors varied greatly with geographic and socio-demographic characteristics across India(3). The ever-increasing burden of cardiovascular disease calls for technological growth to tackle the associated challenges. These needs can be fulfilled, in large part, by the use of electrocardiogram (ECG) for early diagnosis [4]. Considering its non-invasive, readily available and inexpensive nature, it rightly serves as one of the most widely used first-line investigations in the management of heart diseases [5]. These factors both prove and subserve the importance of ECG in cardiovascular medicine when viewed in the context of the rich history of this diagnostic modality, which elegantly stood the tests of time.

The healthcare delivery system in India is broadly grouped under three sectors: the private healthcare sector, the public healthcare centers, and the voluntary health agencies. The Public healthcare system is further divided into primary, secondary, and tertiary healthcare (THC). Primary health care is the first level of contact between communities, individuals, and families with the national health care system. It is provided by a network of primary health centers (PHC), Sub Centres (SC), and Anganwadi centers. It is defined as healthcare based on practical, scientifically sound and socially acceptable methods and technology made universally accessible to individuals and their families at the cost that they can afford [6]. PHCs are operated by medical officers (MOs), nursing, and other paramedical staff who provide both inpatient and outpatient facilities [6]. The primary functions of a medical officer at a PHC are to provide curative care, preventive and promotive care, organize training programs including continuing medical education for the staff at PHC and handle all administrative activities for streamlined functioning of these centers.

PHCs are the first point of contact, especially in rural areas, for patients to seek medical help. Given the knowledge of the course of diagnosis and management of cardiovascular diseases, PHCs may still considerably lack the necessary screening and diagnostic tools that can be used in the early identification of such conditions. The availability of an ECG machine under Project Lifeline in Ahmedabad, Gujarat showed that effective screening in a high-risk population saved 2.90 life years at a cost of 31.07 USD (approximately 2,299.06 INR) per life-year saved, which is below the willingness to pay threshold. Along with the need to increase the availability of such devices, it is also essential to train primary care physicians such that they are better able to interpret ECG readings [7]. For a majority of the underprivileged population, PHCs still remain the first point of contact for health-related care and thus accurate diagnosis and management of all cardiovascular diseases is fundamental in ensuring better prognostic outcomes.

Considering the above factors, the team of World Youth Heart Federation, which was based in Vadodara, Gujarat, undertook a local Pilot Project - CardioGram.Hence, this study was conducted to assess the accessibility, availability, and need for cardiac care with a special focus on ECG at PHCs in the Vadodara district of Gujarat.

## Methods

A cross-sectional pilot study was carried out in the Vadodara District of Gujarat, India between January to March 2022. For administrative purposes, each state is divided into zones which are further divided into districts, and each district is divided into talukas (sub-districts). The District Medical Officer (DMO) oversees all PHCs while each sub-district has at least one PHC headed by an MO. All PHCs present in the Vadodara District were approached to be included in the study. The data collection team initiated the study by contacting the DMO who provided names and contact details for each MO. Following this, individual MOs were contacted via phone call and the basic structure and purpose of the study was explained, and written informed consent was taken. Those MOs who could not be contacted or did not wish to participate in the study were excluded from the study. 34 MOs consented to participate. Hence, 34 out of 42 PHCs were included in the study and were considered for further data collection.

### Study tools and data collection

The data was collected through an online form that was shared with the respective Medical Officer. The form contained 46 questions (37 objective and 9 subjective). The questionnaire was designed to analyze 3 major sections:

- *Accessibility of the PHC* in terms of the closest PHC/THC/CCC (Cardiac Care Centre), distance to the closest PHC/THC/CCC, time taken to reach the closest PHC/THC/CCC
- *Medical Facilities Provided* in terms of availability of ECG, the available first line of treatment for cardiac patients, available drugs for cardiac patients, and management protocol for acute MI patients
- *Staffing of the PHC* in terms of number of currently employed staff, and number of personnel trained to perform an ECGs
- *Need assessment in terms* of the total number of patients admitted in 2019, 2020, 2021, No. of patients visiting the Outpatient Department (OPD) daily, No. of cardiac patients visiting the OPD weekly, Total No. of patients with Hypertension/Diabetes in 2019, 2020, and 2021.

In case of vague and unclear responses on the online form, the MO was contacted via phone call and the response was recorded via interview, and the data was then entered in the form by the data collection team. For questions regarding distance to the closest PHC/CHC/THC/CCC, and time taken to reach the closest PHC/CHC/THC/CCC, the responses were corroborated with data available on Google maps. Any discrepancies in this data were clarified over phone, with the concerned MO.

### Analysis

Data was entered, stored, and analyzed in Google sheets. Descriptive analysis was done in numbers and percentages. Distances were measured in kilometers (km) and Travel times were measured in minutes (min). To assess the need for ECG deployment a Points Based System (PBS) was developed and each PHC is given points on a scale of 0-15 (**Table 1**). PHCs which already have a functional ECG machine and/or where the Medical Officer/Staff are not receptive to utilize the machine were excluded from need assessment.

**Table 1:** Criteria of Point Based System for Need assessment of ECG machine deployment at PHCs.

| Sr. no | Variable | Points given |  |  |  |
| --- | --- | --- | --- | --- | --- |
|  |  | 0 | 1 | 2 | 3 |
| 1 | Distance and time taken to reach closest PHC/ CHC | <30 mins | >30 mins |  |  |
| 2 | Time taken to reach/ Distance from closest tertiary care center/ hospital | <1 hour | >1 hour |  |  |
| 3 | Distance to & Time taken to reach nearest Cardiac Care Hospital | <60 mins | >60 mins |  |  |
| 4 | Population that the PHC/CHC covers (Number) | <30k | >30k |  |  |
| 5 | Are there any trained personnel to take ECG at the PHC/CHC | Yes | No |  |  |
| 6 | If not, are the HCWs willing to learn how to use an ECG Machine | No | Yes |  |  |
| 7 | Are there any trained personnel who can read an ECG | Yes | No |  |  |
| 8 | Are the HCWs willing to learn how to read a normal ECG if we have a training program | No | Yes |  |  |
| 9 | Number of cardiac patients visiting OPD daily | <10 | 10-40 | >40-60 | >60 |
| 10 | Total number of Patients with Hypertension currently registered at the PHC/CHC | <500 | >500 |  |  |
| 11 | Total number of Patients with Diabetes currently registered at the PHC/CHC | <500 | >500 |  |  |
| 12 | Functional ECG Availability | If the answer is yes, the PHC was not considered for deployment |  |  |  |
| 13 | If an ECG machine is deployed at the center, will it be used? | If the answer is no, the PHC was not considered for deployment |  |  |  |
| Classification of PHCs based on Point based Assessment |  |  |  |  |  |
| 0-4 |  | Mild Need |  |  |  |
| 5-9 |  | Moderate Need |  |  |  |
| ≥10 |  | Urgent Need |  |  |  |

## Results

### Accessibility

Distance to the closest next PHC ranged from 0.7 to 40 km with an average of 14.08 km with the time taken to reach ranging from 7 to 60 mins and an average of 22.59 minutes. Distance to the closest tertiary care center to avail tertiary care services ranges from 2 to 40 km with an average of about 37.1 km and travel time ranges from 4 to 120 mins with an average of 62.3 minutes. Distance to the closest Cardiac Care hospital ranges from 2 to 64 km with an average of 55.5 km and travel time ranges from 4 to 95 mins with an average of 85.5 minutes.

### Availability

As summarized in **Table 2**, Only 20 (58.82%) of the 34 PHCs had functional ECG machines available. Among 20 PHCs that had a functional ECG, 11 PHCs had trained Staff Nurses to take ECGs, and 4 PHCs had MOs themselves who would take the ECG. Additionally, 5 PHCs having ECG machines have no trained personnel to either take the ECG or read and interpret it. The possible reasons for underutilization of the ECG machines were found to be lack of motivation (60.6%), insufficient staff (3%), and lack of knowledge (3%) among others. Regarding the availability of medicines at the PHCs, aspirin, beta-blockers were available all in the PHCs (100%), Clopidogrel in 78%, ACE Inhibitors in 96%, Warfarin in 84%, Statins in 81%, Angiotensin Receptor Blockers (ARBs) in 66.66%, Nitrates in 60.6% and Streptokinase in 78.7% PHCs. Functional X-Ray Machines were available in 93.93% PHCs.

**Table 2:** ECG machine availability and its use in PHCs of Vadodara District and Needs Assessment.

| Taluka | PHC name | Name of closest Tertiary Care Center | Name of the closest Cardiac Care Centre/ | Functional ECG Availability | Is ECG Trained Professional Available? | Trained personnel availability for reading an ECG? | Is ECG the first line of approach for chest pain? | Would an ECG machine be used if deployed ? |
| --- | --- | --- | --- | --- | --- | --- | --- | --- |
| Chandod | Prathmik Aarogya Kendra, Chandod | Dhiraj Hospital | SSGH | No | No | No | No | No |
| Sathod | Sathod Prathmik Aarogya Kendra | SSGH | SSGH | No | Yes, Staff Nurse | No | No | No |
| Bhilodiya | Bhilodiya PHC | SSGH | SSGH | No | No | No | No | No |
| Dabhoi | Kayavarohan Prathmik Aarogya Kendra | SSGH | NA | No | No | No | No | Yes |
| Desar | Sandhasal PHC | Parul sevashram hospital | NA | Yes | Yes, Staff Nurse | Yes | No | N/A |
| Karjan | Choranda PHC | SSGH | SSGH | No | No | No | No | Yes |
| Desar | Vejpur PHC | Parul sevashram hospital | NA | No | No | Yes | No | Yes |
| Karjan | Handod PHC | SSGH | SSGH | Yes | Yes, Staff Nurse | No | No | N/A |
| Karjan | Methi PHC | SSGH | SSGH | Yes | Yes, Staff Nurse | Yes | No | N/A |
| Karjan | Rarod PHC | SSGH | SSGH | Yes | Yes, Staff Nurse | Yes | No | N/A |
| Karjan | Valan PHC | SSGH | SSGH | Yes | No | No | No | N/A |
| Padra | Chansad PHC | SSGH | SSGH | No | No | No | No | Yes |
| Padra | Kanzat<br>PHC | SSGH | SSGH | No | No | No | No | Yes |
| Padra | Karkhadi<br>PHC | SSGH | SSGH | No | No | No | No | Yes |
| Padra | Mobha<br>PHC | SSGH | SSGH | Yes | No | No | No | N/A |
| Waghodia | Goraj PHC | Mumi<br>Sevashram | Parul<br>sevashram<br>hospital | Yes | Yes, MO | Yes | No | N/A |
| Waghodia | Asoj PHC | SSGH | Parul<br>sevashram<br>hospital | Yes | Yes, MO | Yes | No | N/A |
| Waghodia | Waghodia<br>PHC | Parul<br>sevashram<br>hospital | Parul<br>sevashram<br>hospital | No | Yes, Staff<br>Nurse | Yes | Yes | Yes |
| Savli | Bhadarva<br>PHC | SSGH | SSGH | Yes | Yes, Staff<br>Nurse | Yes | Yes | N/A |
| Savli | Poicha<br>PHC | SSGH | SSGH | Yes | No | Yes | Yes | N/A |
| Savli | Samalya<br>PHC | SSGH | SSGH | Yes | Yes, Staff<br>Nurse | Yes | Yes | N/A |
| Savli | Vadiya<br>PHC | SSGH | SSGH | Yes | Yes, Staff<br>Nurse | Yes | Yes | N/A |
| Sinor | Sadhli PHC | SSGH | SSGH | Yes | Yes, Staff<br>Nurse | Yes | Yes | N/A |
| Sinor | Simli PHC | SSGH | SSGH | Yes | Yes, Staff<br>Nurse | Yes | Yes | N/A |
| Sinor | Sinor PHC | SSGH | SSGH | Yes | Yes, Staff<br>Nurse | Yes | Yes | N/A |
| Padra | Mujpur<br>PHC | SSGH | SSGH | Yes | Yes, MO | No | No | N/A |
| Padra | Sadhi PHC | SSGH | SSGH | No | No | No | No | Yes |
| Vadodara | Bhayli<br>PHC | GMERS<br>Gotri | GMERS<br>Gotri | Yes | Yes, MO | Yes | No | N/A |
| Vadodara | Angadh<br>PHC | GMERS<br>Gotri | GMERS<br>Gotri | Yes | No | No | No | N/A |
| Vadodara | Sokhda<br>PHC | SSGH | Jamnabai<br>Hospital<br>Chhani | Yes | Yes, Staff<br>Nurse | Yes | No | N/A |
| Vadodara | Kelanpur<br>PHC | SSGH | SSGH | No | Yes, Staff<br>Nurse | Yes | No | Yes |
| Vadodara | Varnama<br>PHC | SSGH | SSGH | No | Yes, Staff<br>Nurse | Yes | No | Yes |

With regards to management of cases of acute chest pain, HCWs at only 2 out of the 33 (6%) PHCs admitted that they took detailed history on presentation. In 96% of the PHCs, the MO handled acute chest pain patients by giving first-line treatment, in the form of Nitrate and Aspirin administration. None of the PHCs treated hypertension or thrombolysed the patient. Even though 57.75% of the PHCs have functional ECG machines, only 24.24% PHCs were using them for managing acute chest pain patients. All of the PHCs referred all the patients of chest pain to a higher center for further management. Only 66.66% of HCWs at PHCs were confident about performing CPR.

### Need Assessment

Healthcare workers (HCWs) in 90.90% of PHCs were willing to learn how to use an ECG machine and HCWs in 78.78% of PHCs were willing to learn how to read ECGs if they received adequate training. When asked about the potential utility of an ECG with adequate training, 66.6% of MOs said that they will use it in High-Risk OPDs, 12% said they will use it in the Regular and High Risk OPDs. Further, 87.87% of the MOs were willing to learn thrombolysis for MI patients by trained cardiologists.

The mean population covered by a PHC was 29978.79. All of the PHCs in total cover a population of 989300 with the maximum population covered by Asoj PHC (56000) in Waghodia Taluka. A mean of 48.84 patients visited the PHC OPD daily. Maximum daily patients load (100/day) were reported to OPDs in Methi and Waghodia PHC daily. Mean Cardiac patient load of all PHCs was 24.7 on a weekly basis. However, Mean cardiac patient load was 170/week at the Sandhasal PHC weekly, where ECG is not being used during first-line treatment of MI even though it is available. The total number of Patients with Hypertension currently registered at the PHCs is 19932 with an average of 622.87 per PHC. Total of 21178 diabetics were registered at all PHCs. Maximum diabetics (5000) and hypertensives (7000) were registered at Sandhasal PHC.

Based on the above factors, Need for ECG deployment was estimated using the Points Based System (PBS) and each PHC was given points on a scale of 0-15. Six PHCs had a score of more than 10 and required urgent deployment of an ECG machine. (**Table 3**)

**Table 3:**
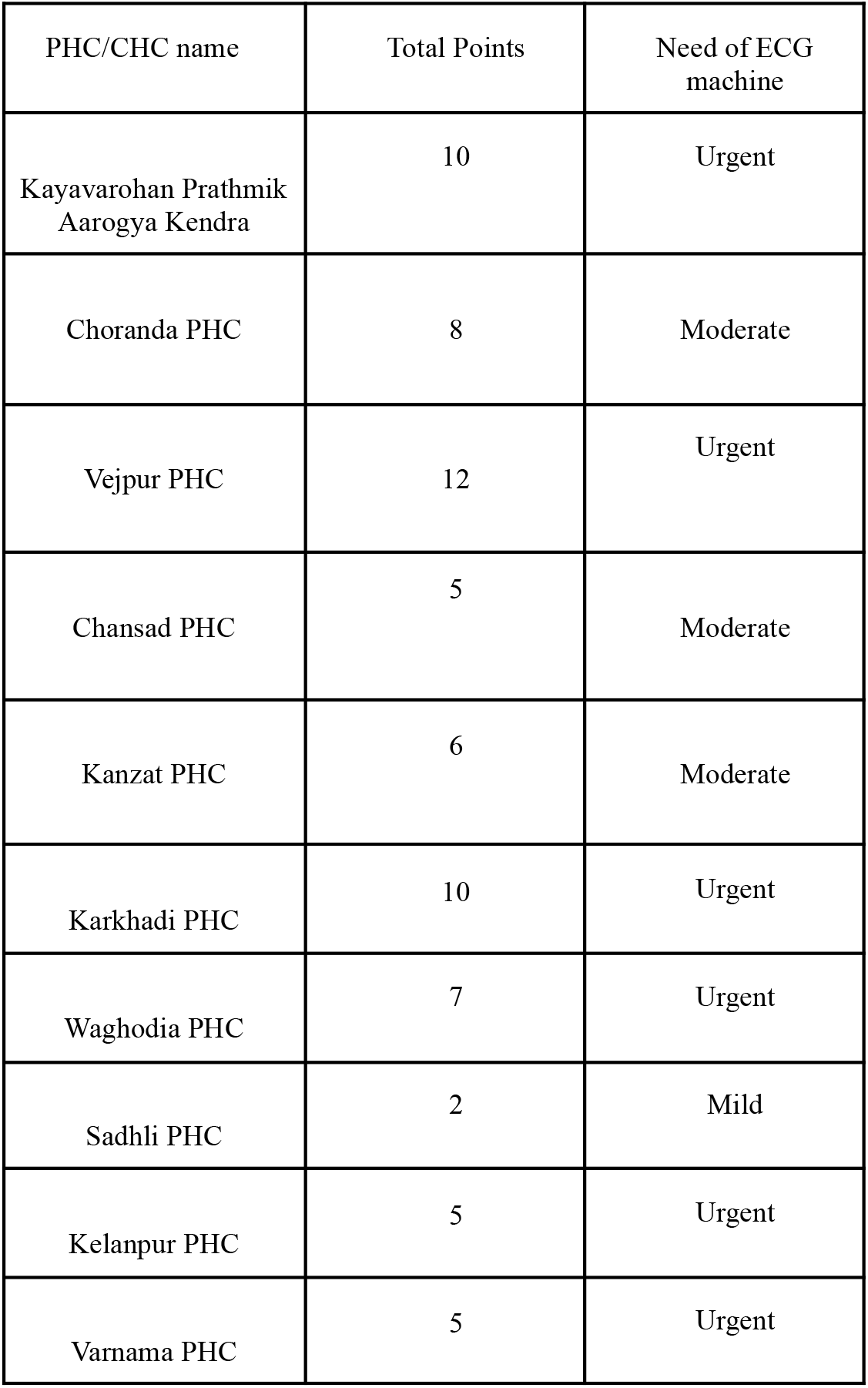
Need assessment of ECG machine deployment at PHCs.

## Discussion

### Summary of results

In this study we assessed the accessibility, availability, and need of cardiac care with special emphasis on ECG availability at PHCs. In the Vadodara district of Gujarat state, even though the travel time to reach a PHC is under 30 mins,the average time taken to reach the nearest THC and CCC from the PHC is over 1 hour. These two travel times combined together exceed the golden time window. Scenario is further worsened by non-availability and non-usage of ECG machines in more than half of the PHCs in the districts, where the Medical Officers are under confident to manage patients with acute chest pain and refer them directly to a higher center without adequate diagnosis and initial management.

### Interpretation and caveats

PHCs make the foundation of our health care system, a place where all essential services should be accessible under one roof. The current scenario seen in the assessment indicates an alarming level of unpreparedness to deal with critical, high-risk, and symptomatic patients, with the closest tertiary and cardiac care centers situated long distances away that would detrimentally affect the survival of the patient and increase morbidity.

The lack of Government funding and resources could explain the low percentage of PHCs having functional ECGs. The Lancet Global Health Commission’s report on PHC financing talks about how it is high time that we not only invest more in financing PHCs but also invest better(8). This patient-centric financing could be achieved by utilizing funds, proper resource allocation, and personnel based on the community’s needs and incentivizing the primary providers. In order to battle the rising incidence of NCDs, the National Programme for Prevention and Control of Cancer, Diabetes, Cardiovascular Diseases, and Stroke (NPCDCS) was launched(9). One of the aims of this program was to increase screening and subsequent early treatment of non-communicable diseases (NCD) like CVDs. Improper implementation of this program along with a lack of awareness regarding it could have resulted in fewerPHCs having functional ECGs - a quick, cost-effective, and non-invasive way of diagnosing CVDs. It is imperative that this gap be filled for proper implementation and coverage of this program.

Currently, many medical colleges lack a curriculum and competency assessment that ensures appropriate learning of ECG interpretation by undergraduate and postgraduate students [10]. Inadequate knowledge about ECG usage and interpretation leads to infrequent administration of ECGs and greater referrals to faraway centers. Competency-based training sessions could help in building confidence and appropriate usage of this diagnostic equipment. Lack of willingness to train was seen in a few PHCs which could arise from the biased preference of patients to be directly referred to advanced tertiary and cardiac care centers instead of continued screening and treatment at PHC [11]. This could be a product of a vicious cycle of PHCs continuously unable to provide optimum care due to improper training leading to patients wanting to be referred directly. The only way to break this chain is sufficient Government funding, providing efficient training sessions, improving equipment, and employing qualified personnel at PHCs.

The above-mentioned results for the availability of functional ECGs do not assess the quality of the machine. Similarly, the results mentioned earlier for the number of personnel who know how to interpret an ECG do not assess the adequacy and/or accuracy of the skill. Furthermore, the data collected for accessibility in minutes does not factor in all factors that could affect it, such as the type of vehicle used and road conditions. Moreover, the availability of drugs present at the PHCs does not indicate the drugs being prescribed correctly to patients with CVD.

### Contextualizing findings

There has been an increase in the burden of CVD, especially in rural India [12]. Integrating screening and treatment for CVD at the primary healthcare level is now a necessity and a missed opportunity [13]. It is seen that morbidity and mortality can be reduced in CVD patients if they get evidence-based treatment, easier access to hospitals, and affordable treatments [14]. All of these can be fulfilled at the primary health care level if functional ECGs are present and HCWs have been trained on how to interpret ECGs effectively and accurately.

A cost-effectiveness analysis done in Ahmedabad, a city in the same state, analyzing the monetary benefits of using a portable ECG for the screening of CVDs among high-risk and symptomatic patients in PHCs showed promising results with 2.9 life years saved at a cost-effectiveness ratio of 2,299.06 INR per life-year saved [7]. The use of portable ECGs at PHCs could justifiably fill the gap uncovered by our need assessment as it makes up only a small proportion of the State Health Expenditure.

The United Nations defines Universal Healthcare as “everyone, everywhere should have access to the health services they need without the risk of financial hardship.” This can be achieved by doing three things - population coverage, covering various services and providing financial security reducing out-of-pocket expenditure. Currently, there exists an imbalance in access to public health in India with various people preferring to be treated at a private hospital leading to excessive out-of-pocket expenditure [15]. Our assessment is in agreement with this. Better funding, allocation of resources, and incentivizing providers could optimize services at PHCs leading to increased access.

### Implications and Future Directions

The findings of this study translate to actionable objectives. As a part of our Pilot Project our NGO raised funds to proquare digital ECGs from manufacturers and donated them to PHCs, who were in urgent need [16]. Our team also conducted training sessions for HCWs at the PHCs for use and interpretation of ECG as well as first line cardiac care training [16]. To scale our local interventions nationally, the Government needs to strengthen infrastructure by financing PHCs via the proper allocation of resources - equipment, and medicines. Secondly, annual competency-based training modules need to be introduced in PHCs via the designing of a “cardiology learning checklist” that includes competencies for interpretation and use of ECGs, designing a proper treatment plan with the correct medication prescribed, correct CPR techniques, etc. A special reward should be given to PHCs that complete this checklist to incentivize providers. The authors plan to scale up the project to all the states of India and also provide more ECGs to PHCs through project Cardiogram initiated by World Youth Heart Federation.

### Strengths and Limitations

To the best of our knowledge, this is the first study done in India that tries to assess the availability of ECGs at the PHC level along with accessibility and need assessment in terms of distance to medical centers, drugs available, administration of ECGs, and following clinical plans like administration of first-line treatment in cases of acute MI. We have also devised a novel Point Based System for the Need assessment of ECG machine deployment at PHCs. However, our findings should be viewed within the context of the study’s limitations. Being a pilot study, the findings are based on one selected district and cannot be generalized to the whole of Gujarat and subsequently, India. Moreover, only 34 out of the 42 PHCs could be included in the study, decreasing the sample size. As data was obtained from the MOs who are in charge of the PHC themselves, there is a high probability of self-reporting along with recall bias.

## Conclusion

Accessibility and availability of cardiac care services, especially ECGs is poor in PHCs of Vadodara district. MOs are unconfident in handling acute MI patients and they are directly referred to Tertiary care centers. There is an urgent need for not only improved services but also efficient training of MOs in order to perform and interpret ECGs as well as effectively manage cases of acute MI.

## Data Availability

All data produced in the present study are available upon reasonable request to the authors

